# Maternal and children’s outcomes for pregnant women with pre-existing multiple long-term conditions: a study protocol of an observational study in the United Kingdom

**DOI:** 10.1101/2022.08.26.22279213

**Authors:** Siang Ing Lee, Holly Hope, Dermot O’Reilly, Lisa Kent, Gillian Santorelli, Anuradhaa Subramanian, Ngawai Moss, Amaya Azcoaga-Lorenzo, Adeniyi Francis Fagbamigbe, Catherine Nelson-Piercy, Christopher Yau, Colin McCowan, Jonathan I Kennedy, Katherine Phillips, Megha Singh, Mohamed Mhereeg, Neil Cockburn, Peter Brocklehurst, Rachel Plachcinski, Richard Riley, Shakila Thangaratinam, Sinead Brophy, Sudasing Pathirannehelage Buddhika Hemali Sudasinghe, Utkarsh Agrawal, Zoe Vowles, Kathryn M Abel, Krishnarajah Nirantharakumar, Mairead Black, Kelly-Ann Eastwood, the MuM-PreDiCT Group

## Abstract

**Introduction:** One in five pregnant women have multiple long-term conditions in the United Kingdom (UK). Studies have shown that maternal multiple long-term conditions are associated with adverse outcomes. This observational study aims to compare maternal and children’s outcome for pregnant women with multiple long-term to those without multiple long-term conditions.

**Methods and analysis:** Pregnant women aged 15 to 49 years old with a conception date between 2000 and 2019 in the UK will be included. The data source will be routine health records from all four UK nations (Clinical Practice Research Datalink [CPRD, England], Secure Anonymised Information Linkage [SAIL, Wales], Scotland routine health records and Northern Ireland Maternity System [NIMATS]), and the Born in Bradford prospective birth cohort.

The exposure of two or more pre-existing, long-term physical or mental health conditions will be defined from a list of health conditions predetermined by women and clinicians. The association of maternal multiple long-term conditions with (i) antenatal, (ii) peripartum, (iii) postnatal and long-term, and (iv) mental health outcomes, for both women and their children will be examined. Outcomes of interest will be guided by a core outcome set.

Comparisons will be made between pregnant women with and without multiple long-term conditions using logistic and Cox regression. Generalised estimating equation will account for the clustering effect of women who had more than one pregnancy episode. Where appropriate, multiple imputation with chained equation will be used for missing data. Federated analysis will be conducted for each dataset and results will be pooled using meta-analysis.

**Ethics and dissemination:** Approval has been obtained from the respective data sources in each UK nation: CPRD: Independent Scientific Advisory Committee (reference: 20_181R); SAIL: Information Governance Review Panel; Scotland: National Health Service Scotland Public Benefit and Privacy Panel for Health and Social Care (HSC-PBPP), The University Teaching and Research Ethics Committee (UTREC) from the University of St Andrews; NIMATS: Honest Broker Service Governance Board; Born in Bradford: Bradford National Health Service Research Ethics Committee (ref 07/H1302/112).

Study findings will be submitted for publications in peer reviewed journals and presented at key conferences for health and social care professionals involved in the care of pregnant women with multiple long-term conditions and their children.

**ARTICLE SUMMARY:** *Strengths and limitations of this study:*

- The study will utilise rich data sources from routine health records from all four UK nations and a birth cohort.
- Beyond examining maternal outcomes, linked mother baby data and the birth cohort data will allow for the exploration of children’s outcomes.
- Key limitations include missing data, misclassification bias due to inaccurate clinical coding and residual confounding.

## INTRODUCTION

One in five pregnant women have two or more long-term physical or mental health conditions prior to pregnancy in the United Kingdom (UK).^1^ In the UK 2016-18 national maternal mortality report, 90% of women who died during or up to a year after pregnancy had multiple health or social problems.^2^ Recent evidence has shown that maternal multiple long-term conditions are associated with adverse outcomes for women and their children, such as severe maternal morbidity and mortality, pre-eclampsia, emergency caesarean birth, preterm birth, and low birth weight.^3-5^

Information on consequences for women with multiple long-term conditions and their children is crucial for women and their health care professionals to make informed decisions on pregnancy care planning. However, there remains a lack of evidence to guide care pathways for pregnant women with multiple long-term conditions.^5 6^

Healthcare is free in the UK and over 98% of the population are registered at a general practice (akin to family practice in other countries).^7^ General practices not only provide primary and community healthcare, but they also serve as the main point of contact for referrals to specialist clinical services and provide the majority of prescribing outside of a hospital setting.^7^ In the UK, pregnant women are recommended to have their booking appointment before 10 weeks gestation.^8^ This is the pregnant woman’s first midwife or doctor appointment, where they undergo health and social care assessment of needs and risks for her pregnancy.^9^ Over 97% of births occur in healthcare settings in England and Wales.^10^ Therefore, routine health records in primary and secondary care in the UK offer a rich data source for observational studies of pregnant women and their children.

This observational study aims to compare outcomes for women with multiple long-term conditions to those without multiple long-term conditions. Outcomes studied will include those for women and their children. Datasets from routine health records from all four UK nations (England, Wales, Scotland, and Northern Ireland) will be used. In addition, the Born in Bradford birth cohort from a deprived, ethnically diverse city in the UK, will also be used.^11^

The four research objectives are to examine the association between maternal pre-existing multiple long-term conditions with: (1) antenatal, (2) peripartum, (3) postnatal and long-term outcomes, and (4) mental health outcomes. The findings from each research objective will be published in a separate paper.

## METHODS AND ANALYSIS

### Study design

This is an observational study using data from routine healthcare records and a prospective birth cohort in the UK.

### Study population and eligibility criteria

The study population will consist of women aged 15-49 years old at conception, with pregnancies beginning between 2000 and 2019 in the UK. Date of conception (pregnancy start date) will be defined as the first day of the last menstrual period or gestational day 0. To ensure sufficient quality data, eligible women must have health records that meet the standard data quality checks as defined by each data source and one year’s worth of health records prior to index pregnancy.

### Data sources

Five population-based data sources (four routine health record datasets and one prospective birth cohort) that will be used are described as follows.

#### (1) Clinical Practice Research Datalink (CPRD), England

CPRD contains anonymised, longitudinal medical records collected during provision of routine healthcare, from participating general practices in the UK; currently 5% of UK general practices contributes to the database.^12^ It includes data on demographics, diagnoses, symptoms, signs, tests and prescriptions.^7^ Linkage to area based deprivation index, Hospital Episodes Statistics and Office for National Statistics death registration data is available for patients whose general practices have consented to the CPRD linkage scheme.^7^

Within CPRD, the CPRD Pregnancy Register is an algorithm that takes information from maternity, antenatal and birth health records from primary care to detect pregnancy episodes and their outcomes.^13^ The Mother Baby Linked data, similarly links women and child records using an algorithm,^14^ will allow for studying the outcomes of children born to mothers with multiple long-term conditions.

CPRD has data for patients from all four UK nations, but analyses using CPRD data will only include English general practices to avoid duplication of patients with datasets from the devolved nations (Wales, Scotland and Northern Ireland).

#### (2) Secure Anonymised Information Linkage (SAIL), Wales

The SAIL databank is a whole population level database in Wales. It is a repository of anonymised health and socio-economic administrative data and provides linkage at an individual level.^15^ It holds data for 4.8 million people and covers 80% of Welsh general practices.^15^ Within SAIL, the National Community Child Health Dataset will be used to detect births and linked to the Welsh Longitudinal General Practice dataset and the Welsh Demographic Service dataset for diagnoses, prescriptions and demographics data respectively.

#### (3) Scotland

A dataset will be created linking the Scottish Maternity Records (SMR02) to data from Hospital Admissions (SMR01), Mental Health Inpatients (SMR04), Accident and Emergency, and the Demography and Death registry. This will cover diagnoses and demographic data for all inpatient stays and day cases for residents in Scotland. The dataset will also be linked to the Prescribing Information System for data on all medications dispensed in the community. Pregnancies will be detected from maternity records or pregnancy-related hospital admissions.

#### (4) Northern Ireland Maternity System (NIMATS)

NIMATS holds demographic and clinical information on mothers and infants.^16^ It captures data relating to the current complete maternity process, and the women’s past medical and obstetric history.^16^ It is a key source for data on birth numbers, interventions, maternal risk factors, birth weights, maternal smoking, body mass index and breastfeeding on discharge.^16^ NIMATS covers all five Health and Social Care Trusts areas across Northern Ireland (11 hospitals providing maternity services in total).^16^ Access to NIMATS is also available to midwives and clerical staff in various community clinics across NI to allow for booking appointments to be recorded.^16^ Pregnancies will be determined from maternity records derived from data recorded at booking appointments (the first antenatal appointment) and hospital admission for childbirth.

#### (5) Born in Bradford

This birth cohort follows over 13,500 children born from around 12,500 mothers at the Bradford Royal Infirmary between March 2007 and June 2011.^17^ Data is collected from pregnancy through childhood and into adult life.^17^ The database consists of over 13,500 pregnancies with biological samples, sociodemographic data, offspring developmental, clinical and education data and is linked to health care records from maternity, primary care (mother and offspring) and hospital admissions.^17^

### Exposure

The exposed group will consist of pregnant women with multiple long-term conditions. We shall define this as two or more long-term physical or mental health conditions that pre-existed before pregnancy. Pregnancy related complications will not be included as they will be studied as outcomes. Multiple long-term conditions will be defined from a list of health conditions previously described in our epidemiological work.^1^ Sensitivity analysis will be performed defining maternal long-term conditions with a different list of health conditions by D’Arcy et al.^18^

Exposure will be ascertained by the presence of diagnostic or prescriptions codes, including Read (to identify exposures in primary care data), International Classification of Disease 10^th^ version (ICD-10, secondary care) and Operating Procedures Codes (OPCS) Classification of Interventions and Procedures.

### Comparator

Pregnant women with no multiple long-term conditions (i.e. no or single long-term conditions) will be the comparator group. Comparisons will be made with the following exposure group:

i. pregnant women with multiple long-term conditions;
ii. pregnant women with increasing counts of long-term health conditions;
iii. pregnant women with different combinations of long-term health conditions.
iv. pregnant women in different health condition clusters (identified from ongoing clustering analyses); and

In addition, we will also compare the outcomes for pregnant women who have mental health conditions as part of their multiple long-term conditions with pregnant women with multiple long-term conditions who do not have mental health conditions.

### Outcomes

The outcomes will be grouped into the following four categories based on the research objectives: (1) antenatal, (2) peripartum, (3) postnatal and long-term outcomes, and (4) mental health outcomes. Examples of outcomes are provided as follows, based on existing core outcome sets for pregnancy and childbirth.^19 20^ The definitive list of outcomes will be confirmed once the development work for a core outcome set for studies of pregnant women with multiple long-term conditions is completed.^21^ Outcomes will be ascertained using Read, ICD-10 and OPCS codes.

#### (1) Antenatal

Antenatal outcomes occur from conception to before the onset of childbirth. Examples for women include miscarriage, gestational hypertension, pre-eclampsia, gestational diabetes, venous thromboembolism and placenta abruption. Examples for children include fetal growth restriction.

#### (2) Peripartum

Peripartum outcomes occur during and immediately after childbirth. This category will also include survival outcomes for women and children. Examples for women include mode of birth (spontaneous vaginal birth, birth with forceps/ ventouse, caesarean birth), postpartum haemorrhage, severe maternal morbidity and maternal death. Examples for children include preterm birth, small for gestational age, stillbirth, perinatal death and neonatal death.

#### (3) Postnatal and long-term

Postnatal outcomes occur in the 42 days after birth.^22^ We will also include perinatal health care utilisation outcomes and long-term outcomes enduring beyond the peripartum and postpartum period. Examples for women include incontinence. Examples for children include congenital anomaly and neurodevelopmental disorders. Examples for health care utilisation include admission to intensive care.

#### (4) Mental health

Mental health outcomes cover the antenatal and postnatal period. Mental health outcomes will be considered up to 12 months after birth. This is to account for possible delay in women presenting to clinicians and reaching a formal diagnosis. Examples include postnatal depression, puerperal psychosis, post-traumatic stress disorder, self-harm and suicide attempts. Children’s mental health and behavioural disorders will also be considered.

### Covariates

Analyses will adjust for the following covariates in a hierarchical manner to explore potential mediating effects. Additional covariates may be added for individual outcomes based on the literature. Where data for antenatal exposures are available (e.g. from NIMATS and Born in Bradford’s booking appointments), additional analyses may be conducted where appropriate.

#### (i) Maternal age

We shall explore whether the association between maternal age and the outcomes are linear. Where this is not the case and to aid clinical interpretability, we will categorise maternal age at conception into 5-yearly age bands.

#### (ii) Parity/gravidity

The variable used will depend on availability in study datasets. Where both variables are available, both will be reported with preference given to *parity* (the number of times a woman gave birth at gestation ≥24 weeks); and sensitivity analysis will be conducted using *gravidity* (the number of times a woman has been pregnant).

#### (iii) Ethnicity

Maternal ethnicity will be categorised based on the variables available in the study datasets: Asian, Black, Mixed, Other and White. Where numbers are too small and risk identifying individuals, such as in NIMATS, we may collapse the categories to White and Non-white.

#### (iv) Social deprivation

The patient level Index of Multiple Deprivation specific to each nation will be used and categorised into quintiles.

#### (v) Body mass index

We shall include the latest available pre-pregnancy body mass index (BMI) for the pregnant women. Where booking data is available before 16 weeks gestation, this will be used (e.g. in NIMATS). BMI will be considered a covariate instead of a health condition. The World Health Organisation’s classification of obesity will be used to categorise BMI: <18.5 kg/m^2^, 18.5 to 24.9 kg/m^2^, 25.0 to 29.9 kg/m^2^, 30.0 to 34.9 kg/m^2^, 35.0 to 39.9 kg/m^2^, and 40+ kg/m^2^.^23^ Categories may be combined where numbers are too small.

#### (vi) Smoking

We shall include the latest available pre-pregnancy smoking status for the pregnant women. Smoking status will be categorised as: non-smoker, ex-smoker, and smoker.

#### (vii) Year (pregnancy start date)

Data quality and clinical guidelines may vary by year. Its effect on outcomes will be accounted for by adjusting for year of conception in the analysis.

### Statistical analysis

Baseline characteristics of the study population and outcomes will be described with summary statistics. Multivariable logistic regression will be performed to estimate the odds ratios for selected study outcomes. Cox regression will be performed for longer-term outcomes. The unit of analysis will be the pregnancy episode.

The covariates will be adjusted for in a hierarchical manner. This is because our prior epidemiological study observed that BMI and smoking may mediate the association between multiple long-term conditions with social deprivation.^1^

A federated analysis approach will be used, each dataset will be analysed separately within the approval of the data access. The effect sizes from the different datasets will then be included in a meta-analysis to produce a summary measure.

Where rare combinations of health conditions and outcomes may lead to identification of an individual or at the prespecified minimum count allowed by each data source, we will suppress the output.

### Pregnant women with more than one pregnancy episode

An individual may have more than one pregnancy over the study period. The pregnancy episodes of the same woman will not be independent of each other. The severity of the exposure variable (pre-existing multiple long-term conditions) may increase in later pregnancy episodes as the pregnant women accumulates more long-term health conditions. If a woman had an adverse pregnancy outcome, she is more at risk of the same adverse outcome in subsequent pregnancy episodes. We shall account for this clustering effect of women with more than one pregnancy episode during the study period using the Generalised Estimating Equation in the regression analyses.

### Multiple pregnancies

The main analysis will be limited to singleton pregnancies. Outcomes for pregnant women with multiple long-term conditions and multiple pregnancies (i.e. twins and higher order pregnancies) will be analysed as a separate cohort.

### Missing data

Where exposure and outcome conditions are identified based on diagnostic codes, the absence of the code will be considered as an absence of the condition. The level and types of missingness of covariates will be reviewed and where appropriate will be addressed with representing missing data as a separate category or multiple imputation with chain equation (MICE). For variables required to compute an outcome, missing values will be imputed using MICE. Example of these variables include birthweight, gestational age and baby’s sex to determine preterm birth and small for gestational age. For each outcome, the statistical analyses will be performed on the imputed datasets and the estimates will be pooled with Rubin’s rule.

### Sensitivity analysis

We shall conduct sensitivity analysis using (i) complete case analysis and (ii) varying definitions of maternal multiple long-term conditions exposure using D’Arcy et al’s core exposure set.^18^

### Patient or user group involvement

The research question was informed by discussions with our patient and public involvement (PPI) advisory group and our PPI co-investigators NM and RP.

The selection of outcomes are guided by our ongoing work developing a core outcome set for studies of pregnant women with multiple long-term conditions, where patients are key stakeholders.^21^

Our PPI advisory group and PPI co-investigators will be involved in interpreting the study findings, producing lay summaries and infographics, and disseminating the study findings through their network.

### Dissemination

Study findings will be submitted for publications in peer reviewed journals and presented at key conferences for health and social care professionals involved in the care of pregnant women with multiple long-term conditions and their children. We will also organise dissemination events to share our findings with the public, service users, clinicians and researchers.

## DISCUSSION

MuM-PreDiCT is a consortium across all four nations of the UK studying multiple long-term conditions in pregnancy. As part of MuM-PreDiCT’s program of work, we outlined the protocol for an observational study of maternal and children’s outcome for pregnant women with multiple long-term conditions, using routine health records and a birth cohort in the UK.

### Comparison with current literature

A recent systematic review found seven observational studies on the association of pre-pregnancy multiple long-term conditions with adverse maternal outcomes.^5^ The review found that pre-pregnancy multiple long-term conditions were associated with severe maternal morbidity, hypertensive disorders of pregnancy, and acute health care use in the perinatal period.^5^ Most studies were conducted in the United States.^5^ Authors of the review commented that many studies included conditions arising in pregnancy in defining multiple long-term conditions, making it difficult to examine the impact of chronic conditions on maternal health.^5^

This proposed study will be based in the UK and will use a broad range of long-term conditions selected by women and clinicians to define multiple long-term conditions. Pregnancy related conditions and complications will be treated as study outcomes and not included in the exposure’s definition. We will also study outcomes across all stages of pregnancy and outcomes for both women and their children.

### Strengths and limitations

This proposed study will utilise routine health records from all four nations of the UK (England, Scotland, Wales and Northern Ireland). The available data sources consist of anonymised patient records from primary and secondary care, community prescription data, and maternity care data from routine booking appointments (first antenatal appointment offered universally and as the gateway to access maternity care in the UK).

Rich data will also be available from a prospective birth cohort from Bradford, an ethnically diverse population in England. Beyond examining maternal outcomes, linked mother baby data and the birth cohort data will allow for the exploration of children’s outcomes. The key strength of this proposed study therefore is the generalisability of study findings to the UK population.

As this is an observational study using anonymised routine health records, key limitations include missing data, misclassification bias due to inaccurate clinical coding and residual confounding.

### Clinical implications

Current obstetric guidelines for pregnant women with medical conditions are focused on specific and single health conditions.^24^ There are currently no guidelines for the management of pregnant women with multiple long-term conditions in the UK. As observed in the systematic review, there is currently a lack of evidence on the consequences of pregnancy for women with multiple long-term conditions.^5^ Our PPI advisory group and preliminary findings from our core outcome set development work have highlighted how women valued having information to help them mentally prepare to face potential adverse pregnancy outcomes.

The output from this study will therefore provide valuable information for women to make informed decision with their clinicians about family planning and their preconception, pregnancy and postpartum care. It will also provide valuable information to guide the future design of care pathway for women with multiple long-term conditions.

## Conclusion

This protocol outlines the study design of an observational study quantifying maternal and children’s outcomes for pregnant women with multiple long-term conditions. The outputs from this study will add to the current body of literature and provide valuable information to help women and their clinicians with their preconception, pregnancy and postpartum care planning.

## Data Availability

In the proposed study, the data that support the findings are available from CPRD, SAIL, Scotland National Health Service Scotland Public Benefit and Privacy Panel for Health and Social Care, NIMATS and Born in Bradford, but restrictions apply to the availability of these data, which were used under license for the current study, and so are not publicly available. Data are however available from the authors upon reasonable request and with permission of CPRD, SAIL, Scotland National Health Service Scotland Public Benefit and Privacy Panel for Health and Social Care, NIMATS and Born in Bradford.

## ABBREVIATIONS

BMI: Body mass index
CPRD: Clinical Practice Research Datalink
ICD-10: International Classification of Disease 10^th^ version
MICE: Multiple imputation with chain equation
NICE: National Institute for Health and Care Excellence
NIMATS: Northern Ireland Maternity System
OPCS: Operating Procedures Codes
PPI: Patient and public involvement
SAIL: Secure Anonymised Information Linkage
UK: United Kingdom

## Ethics approval

### CPRD

CPRD has broad National Research Ethics Service Committee ethics approval for purely observational research using the primary care data and established data linkages. The study has been reviewed and approved by CPRD’s Independent Scientific Advisory Committee (reference: 20_181R).

### SAIL

In accordance with UK Health Research Authority guidance, ethical approval is not mandatory for studies using only anonymised data. The study has been approved by SAIL Information Governance Review Panel.

### Scotland dataset

The study has been approved by the National Health Service Scotland Public Benefit and Privacy Panel for Health and Social Care (HSC-PBPP) and The University Teaching and Research Ethics Committee (UTREC) from the University of St Andrews.

### NIMATS

The study has been approved by the Honest Broker Service Governance Board.

### Born in Bradford

Ethics approval was granted by Bradford National Health Service Research Ethics Committee (ref 07/H1302/112) for the Born in Bradford cohort.

The proposed study is purely observational and will use anonymised research data. The study will not involve participant recruitment. Therefore, consent to participate is not required.

## Consent for publication

This is not applicable as the manuscript is a study protocol. In the proposed study, we will use de-identified study data, therefore consent for publication will not be required.

## Availability of data and materials

This is not applicable as the manuscript is a study protocol. In the proposed study, the data that support the findings are available from CPRD, SAIL, Scotland National Health Service Scotland Public Benefit and Privacy Panel for Health and Social Care, NIMATS and Born in Bradford, but restrictions apply to the availability of these data, which were used under license for the current study, and so are not publicly available. Data are however available from the authors upon reasonable request and with permission of CPRD, SAIL, Scotland National Health Service Scotland Public Benefit and Privacy Panel for Health and Social Care, NIMATS and Born in Bradford.

## Author contributions

SIL – Conceptualisation, funding acquisition, methodology, writing (original draft preparation).

KN, MB, KAE, KMA, DOR - Conceptualisation, funding acquisition, methodology, supervision, writing (review and editing)

HH, GS, AS, NM, AAL, AFF, CNP, CY, CMC, JIK, PB, RP, RR, ST, SB, UA, ZV-Conceptualisation, funding acquisition, methodology, writing (review and editing)

LK, KP, MS, MM, NC, SPBHS - Conceptualisation, methodology, writing (review and editing)

All authors read and approved the manuscript.

## Acknowledgements

Not applicable.

